# Repeat testing for SARS-COV-2: Persistence of viral RNA is common, and clearance is slower in older age groups

**DOI:** 10.1101/2020.08.27.20183483

**Authors:** Paulina Stehlik, Kylie Alcorn, Anna Jones, Sanmarié Schlebusch, Andre Wattiaux, David Henry

**Affiliations:** Evidence-Based Practice Professorial Unit, Gold Coast University Hospital and Health Service, Southport, Queensland, Australia; Institute for Evidence-Based Practice, Bond University, Robina, Queensland, Australia; Infectious Diseases Department, Gold Coast University Hospital and Health Service, Southport, Queensland, Australia; Pathology Queensland, Health Support Queensland, Queensland Health, Queensland, Australia; Queensland Health Forensic and Scientific Services, Brisbane, Queensland, Australia; Gold Coast Public Health Unit, Gold Coast University Hospital and Health Service, Carrara, Queensland, Australia

## Abstract

**OBJECTIVE:** The Novel Coronavirus, or severe acute respiratory syndrome coronavirus 2 (SARS-CoV-2), suppression program has been relatively successful in Queensland, Australia. Initially, it involved extensive community testing and repeat sampling of positive individuals for release from isolation. This enabled study of several characteristics, including persistence of detectable virus and how apparent viral clearance rates varied by age and sex.

**DESIGN:** We conducted an exploratory analysis of Queensland Pathology SARS-CoV-2 reverse transcriptase polymerase chain reaction (RT-PCR) test results. Kaplan Meier analyses were used to estimate median time to apparent viral clearance, and Cox regression to explore the effects of sex and age.

**SETTING AND PARTICIPANTS:** Individuals tested for presence of SARS-CoV-2 in the upper respiratory tract between January 19 and June 4, 2020.

**OUTCOME MEASURES:** Presence of viral RNA detected by RT-PCR.

**RESULTS:** We analysed 97,476 individuals. Median age was 41y (range <1-105y), and 57.2% (95% CI 57.2, 57.2) were female. In total, 958 (0.98%; 95% CI 0.92,1.05) tested positive for SARS-CoV-2. Positivity rates were lower in regional areas than cities, in females (OR 0.80, 95% CI 0.70, 0.91), and in those aged 16y and below (p<0.01, test for trend).

Of the 958 positive individuals, 243 had two or more (maximum 17) additional tests, and 92% (95% CI 88.1, 95.2) remained positive after 10 days (maximum 72 days) after the initial result.

Median time to apparent viral clearance was longer in those 65y and over compared to those under 65y (29 v 43 days, HR 1.85; 95% CI 1.17, 2.90), and was unaffected by sex (HR 0.93; 95% CI 0.66, 1.30).

**CONCLUSIONS:** Females and those 16y and under were less likely to test positive for SARS-CoV-2. Detectable RNA may persist for long periods, negating the value of repeat testing for declaring individuals free of infection. Viral clearance rates appear lower in those over 65y of age compared with younger individuals.

“The known”

- Early in the COVID-19 pandemic, 2 negative RT-PCR swabs were used to achieve negative status in infected individuals
- There are few published data on the patterns of results seen with repeat testing.

“The new”

- We analysed data from a large cohort of people tested for viral RNA in Queensland, Australia.
- We found that females and those 16 y and under were less likely to test positive.
- Viral RNA was detectable for up to 72 days, with >90% testing positive for more than 10 days.
- Viral clearance was slower in those over the age of 65.

“The implications”

- Our findings support that there is likely to be little value in repeat RT-PCR testing to declare individuals free from infection.

## BACKGROUND/RATIONALE

The first Australian cases of infection with severe acute respiratory syndrome coronavirus 2 (SARS-CoV-2) were reported in January 2020.(1) During the initial phase the peak daily infection rate was in late March 2020 and by the end of June 2020 there had been around 8000 cases and 104 deaths.(2) Initially, the majority of cases were acquired outside the country rather than by local transmission.(3) The cumulative incidence rates (June 2020) of around 400/million, and mortality of 4/million, were towards the lower end of the rates that have been experienced in other high-income countries, although these are rising quickly with recent outbreaks in Victoria and New South Wales.(3) Rates of infection remain relatively low in Queensland.(3)

In Queensland, the criteria for testing individuals for SARS-CoV-2 changed during the pandemic. Initially, to be tested in Queensland people were required to meet both epidemiological (return from a high-risk country), and clinical criteria (suggestive symptoms). With progression of the pandemic, testing criteria were modified to clinical criteria only (details provided below).

The rollout of a comprehensive testing program and the availability of data from repeated within-subject testing carried out in the initial stages of the Coronavirus Disease 2019 (COVID-19) pandemic provided an opportunity to conduct an exploratory study to address several questions. We investigated population testing rates and how they varied over time. We estimated the proportions of individuals who returned positive tests and how these varied with location age and sex. We also estimated apparent rates of clearance of viral RNA from the upper respiratory tract of subjects with repeated tests, and the extent to these varied with age and sex.

## METHODS

### STUDY DESIGN

Our study sample consisted of individuals who underwent a swab test for SARS-CoV-2 processed in Queensland Health public laboratories between Jan 10^th^ and June 4^th^, 2020. The procedure involved sampling from the nasopharynx or oropharynx using one or two swabs and identification of SARS-CoV-2 RNA was by reverse transcriptase polymerase chain reaction (RT PCR). Rarely, RT PCR of sputum samples may have been used for diagnosis.

The criteria for testing individuals evolved over time. Initially, to be tested, people were required to meet both epidemiological and clinical criteria. The epidemiological criteria were dependent on travel to high risk areas or contact with confirmed cases. As the pandemic spread, the epidemiological criteria expanded to include all international travel. Enhanced testing began in certain areas of Queensland at the beginning of April, which meant that people with no known epidemiological risk could be tested if they met the clinical criteria alone. By the end of April 2020, this enhanced testing was state-wide. Prior to late March release from isolation was based on days since illness onset, symptoms resolution and two swabs 24 hours apart that were negative for SARS-CoV-2. These criteria subsequently changed to release from isolation based on symptoms and duration alone (72 hours since symptom resolution and 10 days since illness onset or discharge from hospital). Exceptions were healthcare and aged care facility workers, who continued to require two swabs in addition to standard requirements for release. This was changed at the beginning of June 2020 to only include individuals who had underlying conditions that made it difficult to define symptom resolution, or who were immunocompromised.

### DATA SOURCES

Data on the results of swab testing for viral RNA were accessed from AUSLAB, a centralized laboratory information system for all pathology results from the Queensland Health public pathology system, including tests ordered from public hospitals, community centres, correctional centres, and private practitioners using the public health pathology services including those referred to Queensland Pathology from other states for processing. AUSLAB dataset do not contain test results processed by private laboratories outside of the pubic pathology system.

All analyses were performed using de-identified data. Clinical information and the reasons for testing were not available. An exemption from full research ethics review was granted by the Gold Coast Health Human Research Ethics Committee (LNR/2020/QGC/63045).

### VARIABLES

Each row of the AUSLAB data represented a unique test result and contained the following variables:

- Unique encrypted individual identifier.
- Date of sample collection.
- Time of sample collection.
- Facility where the test was ordered.
- Age of individual.
- Sex of individual.
- Laboratory number. Multiple results from the same sample were assigned to a single laboratory number.
- Whether SARS-CoV-2 RNA was detected in the sample.

We added an additional geographic variable based on the postcode of the facility where the first test was collected. This was categorized using the definitions of the Queensland Government Statistician’s Office as: Very Remote or Remote, Outer Regional, Inner Regional, and Major City.(4)

### DEFINING INDIVIDUALS’ INFECTION STATUS

The following categorizations were used:

- Individuals with a negative test that was not repeated were classified as negative
- Individuals with a positive test that was not repeated were classified as positive
- In the case of multiple tests on different days individuals were considered to have a negative status until they received a single positive test result and then they were considered positive.
- When both a positive and negative result were obtained on the same day the individual was considered positive.
- Once positive, individuals needed to achieve two consecutive negative test results on samples collected at least 24 hours apart to be assigned a ‘negative status’.
- After a positive result we defined the number of days to negative status as the number of days to the first of the two negative tests.

Data cleaning and classification was performed in Python (Python Software Foundation, version 3.7). Full description of dataset structure, outline of logic, and code are provided on our Open Science Framework (OSF) project page.(5)

### DATA ANALYSIS

Our analyses were mainly descriptive. We wished to determine testing rates and how they varied geographically, what proportion of individuals returned positive tests and how this varied with age and sex, the apparent rates of clearance of viral RNA from the upper respiratory tract and the extent to which this varied with age and sex.

Descriptive statistics are expressed as means or medians (as appropriate) with ranges, and proportions with 95% confidence intervals. We measured associations by Odds Ratios with 95% confidence intervals, used Kaplan Meier analyses to estimate time to negative status and Cox regression analyses to explore the effects of sex and age on apparent clearance rates (< 65y v ≥ 65y). Trend lines were generated using smoothed conditional means; generalized additive model was used for all tests, and loess method was used for positive test results.

All analyses were conducted in R (R Foundation; version 3.6.3). Further details, including methods used to generate heat maps for displaying repeated observations are provided on our OSF page.(5)

## RESULTS

In total, 103,984 swabs were obtained from 97,478 individuals and tested for the presence of SARS-CoV-2 during the 146-day study period. This represented 49.8% (n = 208,758) of all tests conducted in Queensland and 88.96% of all positive cases (n = 1,060).(6)

Two individuals (with one test each) were removed from the analysis due to missing sample collection date and an invalid test result, leaving 97,476 individuals in our analysis. The median age of all individuals who were tested was 41y, and 57.2% (95% CI 57.2, 57.2) were female (Table 1).

**TABLE 1:** DEMOGRAPHIC DATA ON TESTED INDIVIDUALS GROUPED BY RESULTS (10 JAN – 04 JUNE 2020)

| Name | All Individuals n (%) | 95% CI | Negative individuals (%) | 95% CI | Positive individuals (%) | 95% CI |
| --- | --- | --- | --- | --- | --- | --- |
| **Female** | 55708 (57.15) | 57.15, 57.15 | 55212 (57.2) | 57.2, 57.2 | 496 (51.77) | 48.54, 55.14 |
| **Male** | 41594 (42.67) | 42.67, 42.67 | 41132 (42.62) | 42.62, 42.62 | 462 (48.23) | 44.99, 51.59 |
| **Not reported** | 174 (0.18) | 0.18, 0.18 | 174 (0.18) | 0.18, 0.18 | 0 (0) | 0, 3.36 |
| **Median Age** | 41 | 0, 105* | 41 | 0, 105* | 45 | 0, 88* |
| **16 and under** | 11262 (11.55) | 11.27, 11.84 | 11239 (11.64) | 11.64, 11.65 | 23 (2.4) | 0, 5.08 |
| **17 to 64** | 70811 (72.64) | 72.36, 72.93 | 70063 (72.59) | 72.59, 72.59 | 748 (78.08) | 75.57, 80.76 |
| **65 and over** | 15399 (15.8) | 15.52, 16.08 | 15212 (15.76) | 15.76, 15.76 | 187 (19.52) | 17.01, 22.2 |
| **Not reported** | 4 (0) | 0, 0.29 | 4 (0) | 0, 0.01 | 0 (0) | 0, 2.68 |
| **16 and under** | 11262 (11.55) | 11.27, 11.84 | 11239 (11.64) | 11.64, 11.65 | 23 (2.4) | 0, 5.08 |
| **17 to 64** | 70811 (72.64) | 72.36, 72.93 | 70063 (72.59) | 72.59, 72.59 | 748 (78.08) | 75.57, 80.76 |
| **65 and over** | 15399 (15.8) | 15.52, 16.08 | 15212 (15.76) | 15.76, 15.76 | 187 (19.52) | 17.01, 22.2 |
| **Not reported** | 4 (0) | 0, 0.29 | 4 (0) | 0, 0.01 | 0 (0) | 0, 2.68 |

\* Range

### TESTING AND POSITIVITY RATES BY AGE AND SEX

Nine hundred fifty-eight individuals (0.98%; 95% CI 0.92,1.05) tested positive for SARS-CoV-2. The median age of those whose swabs tested positive was higher than that of the whole group (45y), and they were less likely than the whole group to be female (51.8%; 95% CI 48.5, 55.1) (Table 1).

The odds of a positive test increased with age. Compared with those aged 16 y and under the Odds Ratio (OR) for a positive test in those aged 17 to 64 y was 5.2 (3.4, 8.1), and was 6.0 (4.0, 9.5) in those aged 65 y and above (P < 0.01 χ^2^ test for trend across 3 age groups). Females had a lower odds of a positive test than males (OR 0.80, 95% CI 0.70, 0.91).

### TESTING AND POSITIVITY RATES BY REGION

The test positivity rate was higher in cities than in regional areas (Table 2) though the highest rate was seen in a geographically uncategorized group. These included samples collected by private practitioners (positivity rate 3.6%, 95% CI 2.9, 4.4) and a pathology service in the border region with New South Wales (1.9%, 95% CI 1.3, 2.9).

**TABLE 2:** RATE OF TEST POSITIVITY FOR SARS-COV-2 BY LEVEL OF REMOTENESS IN QUEENSLAND* AUSTRALIA

| **Area** | **Individuals tested** | **Individuals who tested positive** | **Percentage positive tests (95% CI)** |
| --- | --- | --- | --- |
| **Unknown** | 3599 | 106 | 2.9 (2.4, 3.6) |
| **Private Practitioner** | 2291 | 82 | 3.58 (2.87, 4.44) |
| **Northern NSW boarder pathology lab\*\*** | 1187 | 23 | 1.94 (1.26, 2.94) |
| **Major Cities** | 62726 | 726 | 1.2 (1.1, 1.2) |
| **Inner Regional** | 13332 | 71 | 0.5 (0.4, 0.7) |
| **Outer Regional** | 15009 | 52 | 0.3 (0.3, 0.5) |
| **Remote and very Remote** | 2810 | 3 | 0.1 (0, 0.3) |

\* Data processed by Queensland Public Pathology contains some NSW data where it had been referred to Queensland Pathology for processing.
\*\* Intentionally unnamed.

### SARS-COV-2 TESTING AND POSITIVE RESULTS OVER TIME

Both the numbers of individuals tested and the numbers in whom SARS-CoV-2 was detected peaked in the second half of March 2020 (Figure 1). Thereafter, there was a decline in testing with a nadir in early April followed by a continuing rise until the end of the study period in early June 2020. At the time of writing there has been no second peak in the number of detections in Queensland and case numbers remain low.

**FIGURE 1:**
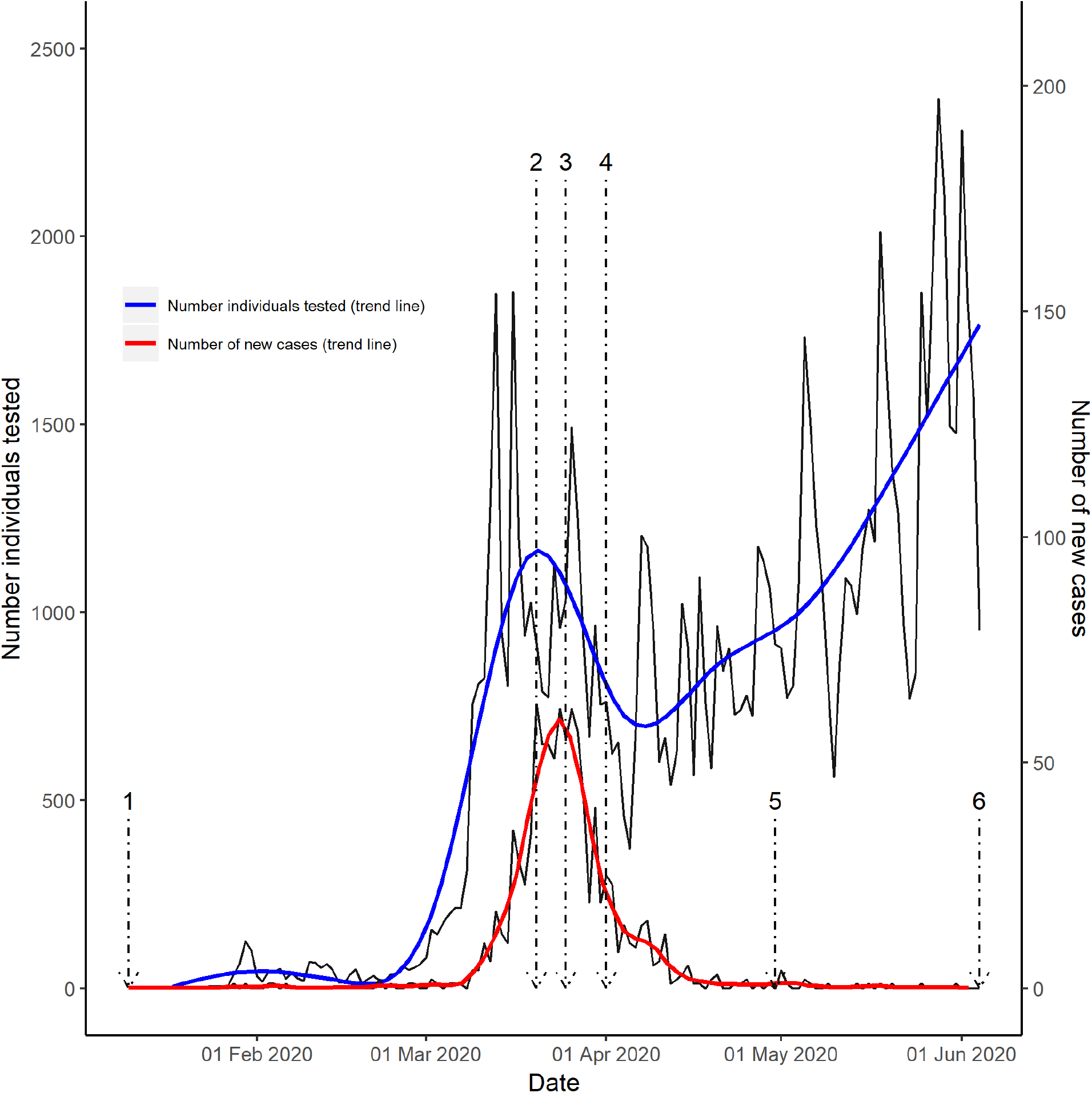
SARS-COV-2 TESTING* AND NUMBERS OF POSITIVE CASES OVER TIME, ANNOTATED WITH DATES OF KEY CHANGES IN TESTING POLICIES IN QUEENSLAND, AUSTRALIA. 1 = (10 Jan) People were required to meet both epidemiological and clinical criteria to be eligible for testing. The epidemiological criteria were travel to high risk areas or contact with confirmed cases. Over time the epidemiological criteria expanded to include an increasing list of countries until all international travel was included. Release from isolation was based on days since illness onset, symptoms resolution AND two consecutive negative tests > 24 hours apart. 2 = (20 March) Australian international borders close. (21 March) Release from isolation criteria changed to symptoms and duration alone (required 72 hours of symptom resolution and 10 days since illness onset or discharge from hospital) except for health care and aged workers. 3 = (25 March) Queensland border closure 4 = (Start of April) Enhanced testing began in certain areas of Queensland. People with no known epidemiological risk could be tested if they met the clinical criteria alone. 5 = (End of April) Enhanced testing implemented across Queensland 6 = (4 Jun) The requirement of swabs for release from isolation for people in high risk settings was removed. Only immunocompromised people required clearance swabs after this date. *Repeat tests after first positive not included. We used generalized additive model for “all tests” (blue line), and loess (Local Polynomial Regression Fitting) for “positive test results” (red line) to generate trend lines.

### RESULTS SEEN WITH REPEAT TESTING

Of the 947 individuals who tested positive, 317 (33.1%; 95% CI 30.1, 36.2) had repeat tests and 243 had two or more repeat tests (25.4; 95% CI 22.6, 28.2). The largest number of repeat days of tests in any individual was 17.

Of the 243 individuals with initially positive results, and at least two repeat tests, 147 (60.5%; 95% CI 54.0, 66.7) were documented as achieving negative status (2 negative results at least 24 hours apart) during the observation period. Most were in the 17-64 y age group and females were more likely to be documented to reach negative status than males OR 1.7 (95% CI 1.0, 2.9) (Supplementary File Table 1).

As is apparent in Figure 2, there was considerable variability in the patterns of within-subject repeat test results. Of 243 individuals who received two or more repeat tests after an initial positive result, the 224 (92.2% 95% CI 88.1, 95.2) had one or more additional positive results after 10 days and up to 72 days after the initial test.

**FIGURE 2:**
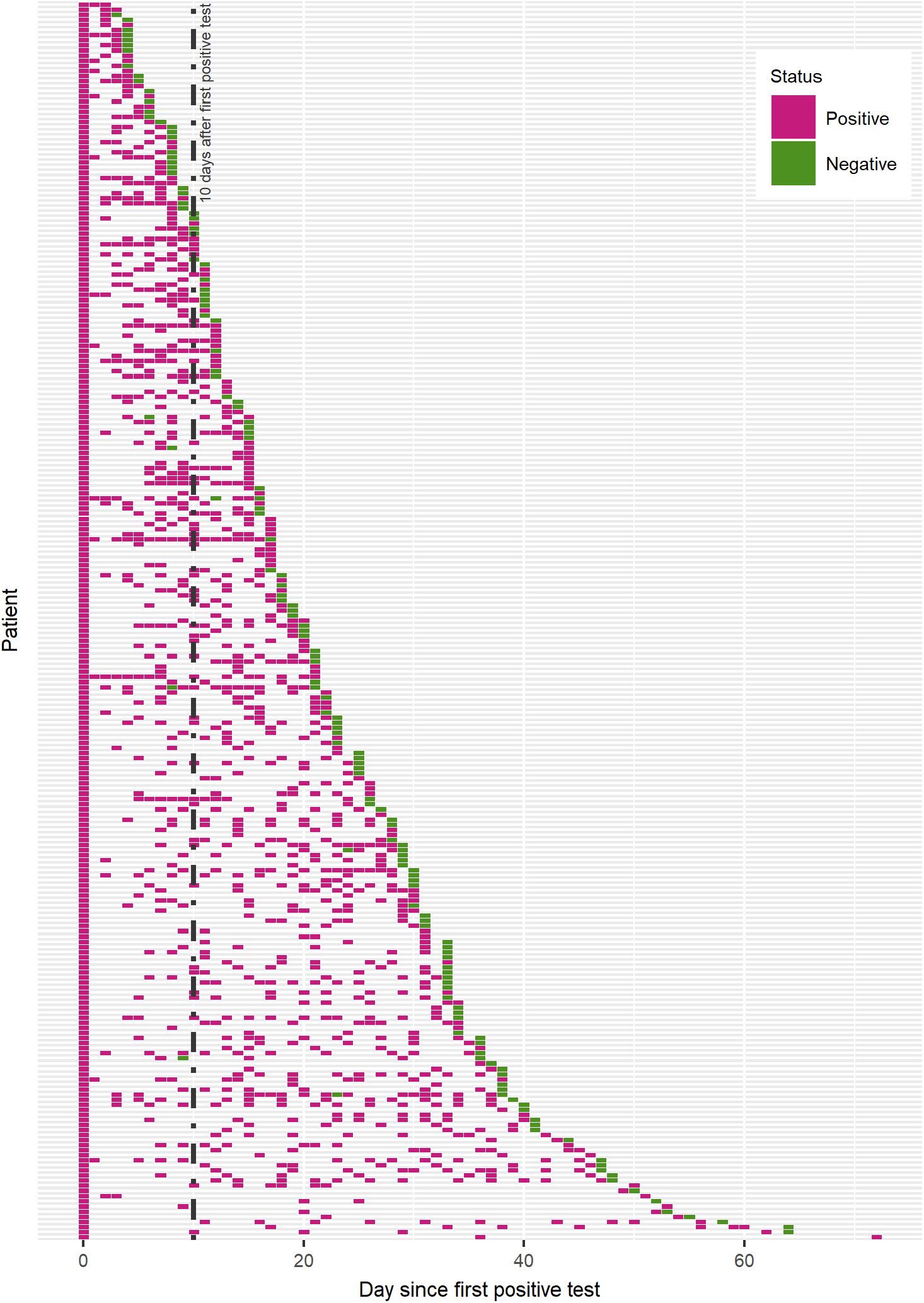
CATEGORICAL HEAT MAP OF TESTING RESULTS IN SUBJECTS WHO TESTED POSITIVE FOR SARS-COV-2 There was considerable variability in the patterns of within-subject repeat test results after an initial positive PCR test. Of 243 individuals who received two or more repeat tests 224 (92.2%) had one or more additional positive results after 10 days and up to 72 days after the initial test. After achieving negative status, 7 out of 147 (4.8%) had a subsequent positive test. Six of these were men.

After achieving negative status, 7 out of 147 (4.8% 95% CI 1.9, 9.6) had a subsequent positive test. Six of these were men.

### APPARENT VIRAL CLEARANCE RATES

The Kaplan Meier analysis of positive individuals who achieved negative status is illustrated in Figure 3. The median time to clearance was 20 days. We repeated this analysis on data from all individuals who had an initially positive test, irrespective of whether they achieved negative status (2 consecutive negative results). The estimated median time to viral clearance was 11 days longer (Figure 4)

**FIGURE 3:**
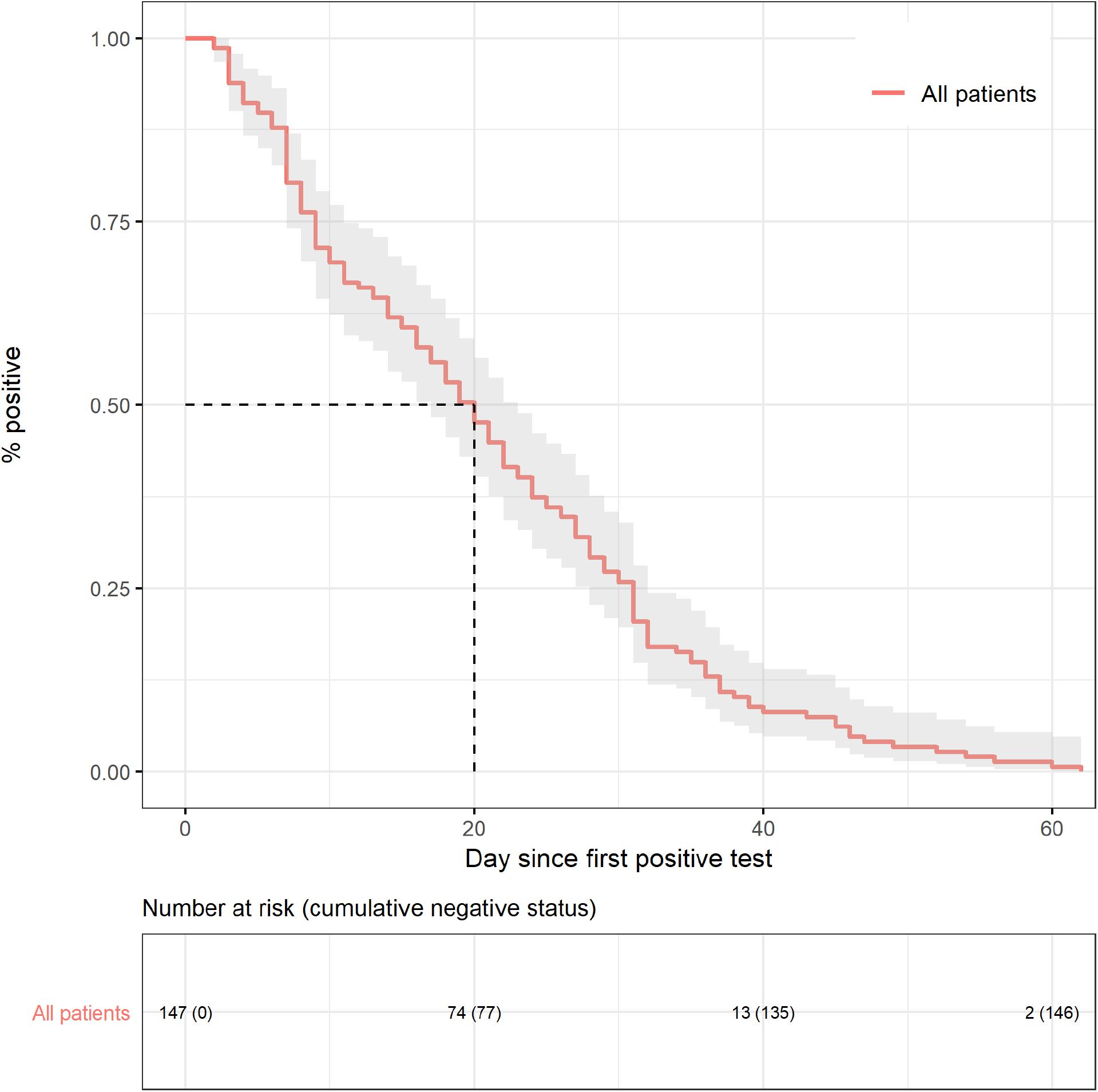
KAPLAN MEIER ANALYSIS OF VIRUS CLEARANCE RESTRICTED TO THOSE SUBJECTS WHO ACHIEVED NEGATIVE STATUS DURING FOLLOW-UP.

**FIGURE 4:**
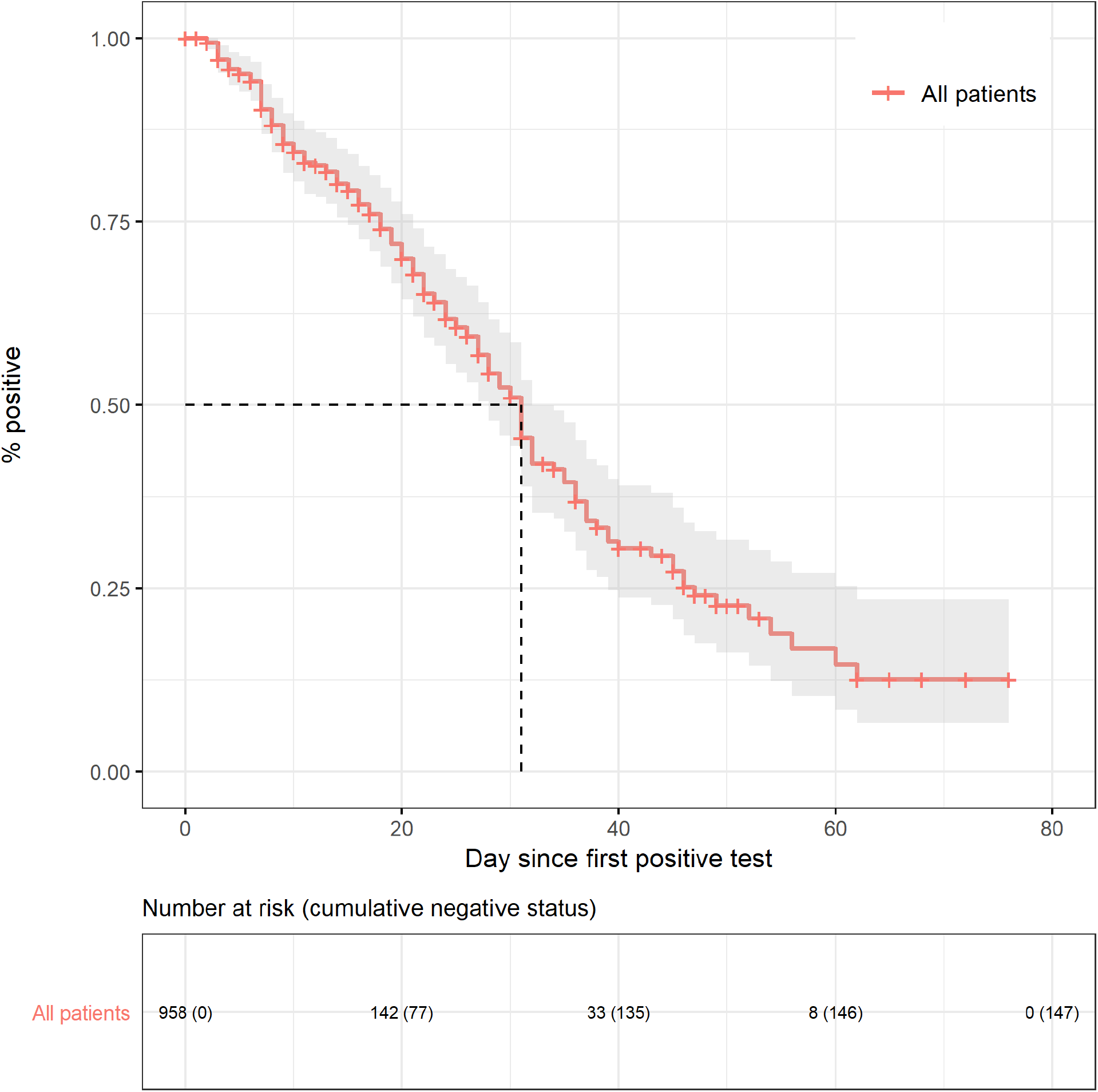
KAPLAN MEIER ANALYSIS OF VIRUS CLEARANCE IN ALL SUBJECTS WHO INITIALLY TESTED POSITIVE FOR SARS-COV-2.

We performed Kaplan Meier and Cox Regression analyses to estimate the effects of age and sex on the apparent clearance rate of the virus in all subjects with an initially positive result. Using negative status as the endpoint we found no effect of sex on rates of clearance (female v male: HR 0.93; 95% CI 0.66, 1.3) but a higher clearance rate in subjects under 65 years (compared with those 65 y and above: HR 1.82 (95% CI 1.17, 2.93)) (Figure 5 and Figure 6).

**FIGURE 5:**
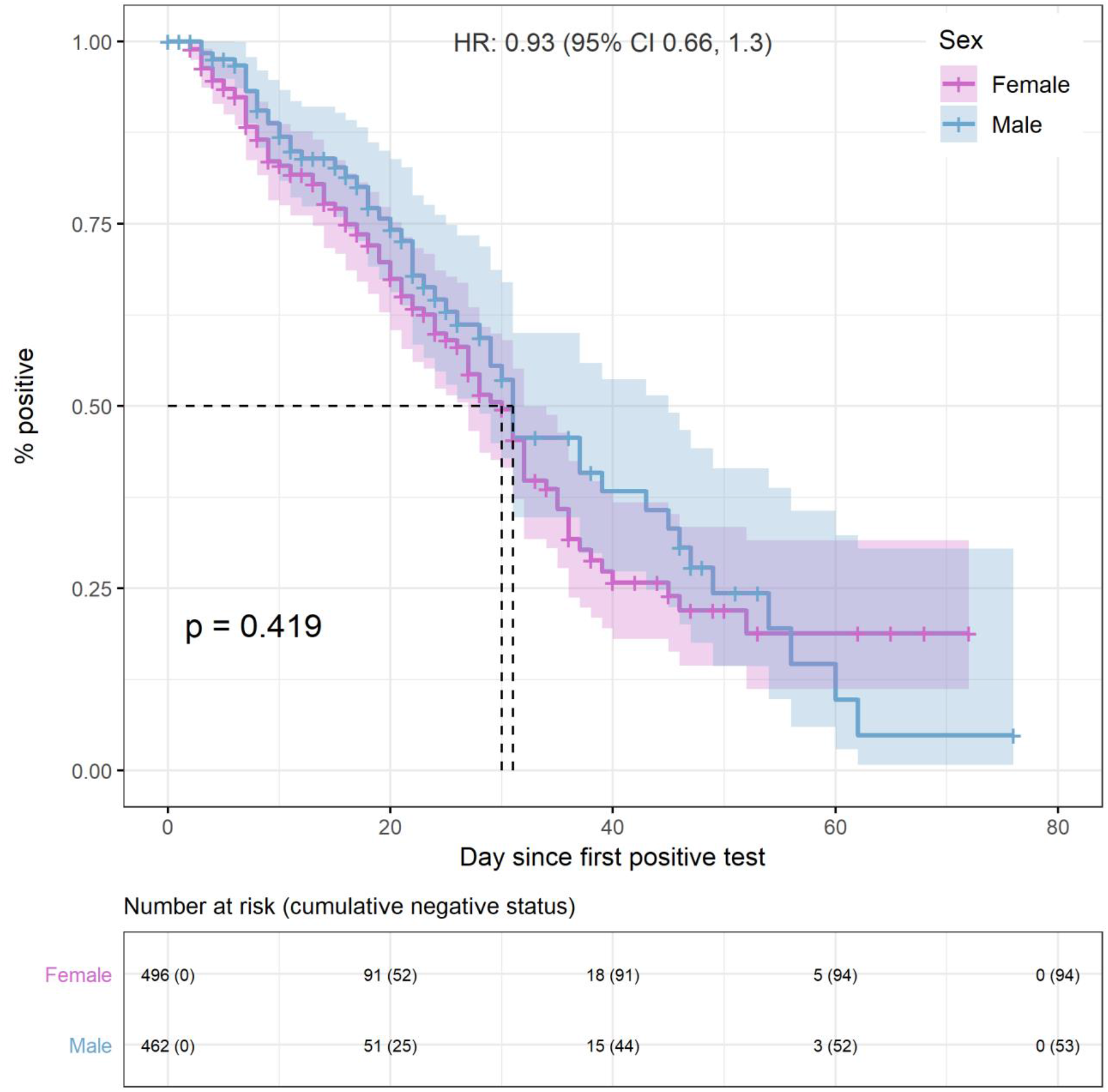
KAPLAN MEIER ANALYSES OF VIRUS CLEARANCE IN ALL SUBJECTS WHO INITIALLY TESTED POSITIVE FOR SARS-COV-2, GROUPED BY SEX. There was no difference in median time to negative status in females v males (30 v 31 days, Hazard Ratio 0.93; 95% CI 0.66, 1.30, p-value = 0.419). Confidence bands were generated using Cox proportional hazards regression model with Efron approximation using the coxph function in R in both panels.

**FIGURE 6:**
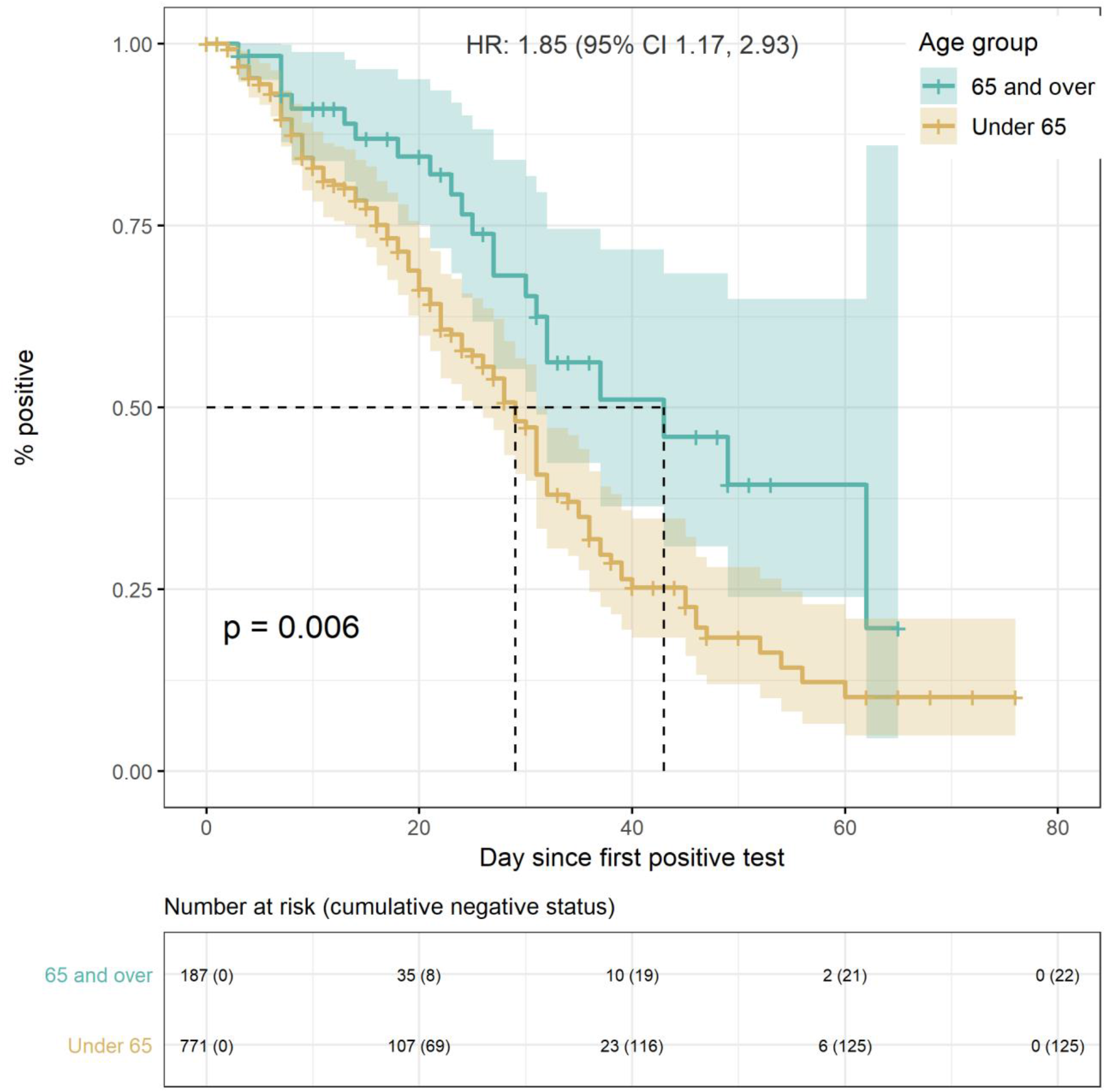
KAPLAN MEIER ANALYSES OF VIRUS CLEARANCE IN ALL SUBJECTS WHO INITIALLY TESTED POSITIVE FOR SARS-COV-2, GROUPED BY AGE. Time to achievement of negative status was shorter in subjects under 65y (median 29 days) compared with older subjects (median 43 days). The Hazard Ratio was 1.85 (95% CI 1.17, 2.93, p-value = 0.006). Confidence bands were generated using Cox proportional hazards regression model with Efron approximation using the coxph function in R in both panels.

## DISCUSSION

Our findings indicate that viral RNA remain in individuals for long periods of time, and that this is even longer in those over the age of 65. We also found differences in positivity rates with lower positivity rates amongst females, those under the age of 16, and in non-urban areas.

None of our findings are novel, but together they highlight some relevant features of the evolution of the infection in Queensland and the patterns of results and viral clearance rates seen with repeat testing. As such, they have local and potentially wider relevance.

At a policy level, the results show how frequently positive swabs were seen 10 or more days after infection; in these results this was documented for up to 72 days. Over 90% of individuals who received two or more repeat tests after an initial positive result had one or more additional positive results after 10 days. This confirms reports of persistence of the viral RNA in airways of some infected individuals. This has been observed in bronchial secretions rather than throat swabs and is not considered indicative of continued viral replication and potential for transmission.(7, 8) Rather, these are likely to be inactive fragments of virus, sometimes called ‘viral litter’.(8) Furthermore, all patients except health and aged care workers, were released from isolation from 21^st^ March onwards based on symptoms and duration alone and local transmission continued to decline to zero. In addition, evidence suggests that viral shedding after 8-10 days from symptom onset are unlikely to be infective.(7) Our findings support the change in policy that no longer mandates repeated testing to demonstrate negative status.

Higher test positivity rates were seen in urban (Major City) than in rural areas of Queensland. There are two possible causes – one is increased rates of transmission due to higher population densities and greater opportunities for person to person contact. The second and more likely is the high proportion of Australian infections that were identified in overseas travellers during the early stages of the pandemic. Tourists enter the state via major centres and tend to visit urban locations rather than remote areas. In addition, after introduction of mandatory quarantine, many were quarantined in city hotels and tested there.

We found a relationship between test positivity rates and age group, with a positive test for trend in the OR across three age groups. The finding of lower rates in youth (aged 16 years or less) has been commented on.(9, 10) This in part may reflect their lower susceptibility to infection. However, travellers contributed to the higher positivity rates early in the pandemic and were predominantly adults, which may have biased our finding.

The odds of finding an initial positive test were 20% lower in females than males. Several studies have suggested that male sex is associated with more severe Covid-19 illness.(11) However, with adjustment for age we found no difference in viral clearance rates between males and females. In contrast, the average rate of achieving negative status was 85% higher in those under 65y of age when compared with those 65y and above. This apparent impairment of elimination of the Coronavirus with advancing age has been reported previously.(12) The magnitude of the effect measured here may have clinical significance. Rates of hospitalisation, admission to intensive care and mortality have been shown to be elevated in older subjects.(13) Our observation of slower viral clearance suggests a decline in immune function related directly to age and/or co-existing disease.

The apparent rates of clearance we observed in this study were low. For instance, in the full population of individuals with initially positive tests the median time to clearance of viral RNA was around 30 days. This dropped to 20 days when analysis was confined to those who achieved negative test status (the first of 2 negative tests separated by at least 24 hours). In comparison, in an early trial of hydroxychloroquine Tang *et al* measured a median time to clearance of 8 days in the control group(14), and Cai *et al* measured a median time to clearance of 11 days in the control arm of a study of Favipiravir.(15) These studies were conducted in hospital with close monitoring of patients and frequent measurements of viral RNA. In contrast, in a retrospective cohort study Kim *et al* measured median clearance times of 21 and 28 days in patients treated with lopinavir/ ritonavir and hydroxychloroquine, respectively.(12) These latter values are close to those we observed. The intermittent testing and incomplete follow-up of individuals in the current study probably led to overestimation of the median clearance times in our analyses. However, we don’t think that this will have introduced a systematic bias in the assessment of the effects of age and sex.

Overall, around 1% of the tests reported here were positive for SARS-CoV-2, although this was weighted by the higher positivity rates seen early in the pandemic. The positivity rates were at, or close to, zero by the end of the study period. Testing rates fell after the initial peak as criteria for testing required a history of overseas travel and the number of incoming travellers declined in late March. Enhanced testing started and rose in April, which explains the subsequent rising curve (Figure 1).

It is important to recognize that this study was descriptive and opportunistic. The timing of collection of samples, particularly repeat swabs, was neither standardized nor consistent. The latter reflected the exploratory nature of testing during the early stages of a pandemic involving a novel virus. We did not have either the indications for testing for SARS-CoV-2, nor any clinical details of the individuals who tested positive. The latter would require linkage of the pathology data to primary care and hospital electronic medical records health administrative and cause of death data. Such comprehensive linkage is not currently widely available in Australia.(16) In addition, our sample of test data may not have been completely representative of the state of Queensland as it does not include results for all tests performed at private pathology laboratories. This bias in the sampling frame may have led to overestimation of the overall positivity rate and the variations we saw between the regions. However, we think it unlikely that sampling bias has distorted the evolving pattern of the pandemic or the conclusions that were derived from analyses of repeated testing for SARS-CoV-2.

## CONCLUSIONS

Despite lack of information on clinical indications for testing, and inconsistent patterns of retesting following positive results, we saw some results of possible importance. Detectable RNA may persist for long periods and negate the value of repeat testing as a basis of declaring individuals free of infection. The probability of testing positive for SARS-CoV-2 in our cohort was lower in those 16 years and under and in females. Viral clearance rates appear to lower in those over 65 years of age compared with younger individuals.

## Data Availability

Original data not available due to privacy; analysis code, description of data structure available at our OSF webpage.

https://osf.io/habvk/

## SUPPLEMENTARY FILES

**TABLE 1.**

| **Name** | **Positive individuals with 2 or more swabs after first positive (%) (95% CI)** | **Positive individuals who achieved a negative status (2 neg tests) (%) (95% CI)** | **Positive individuals with status change from negative back to positive (%) (95% CI)** |
| --- | --- | --- | --- |
| **SEX** |  |  |  |
| **Female** | 143 (58.8) (52.7, 65.3) | 94 (63.9) (56.5, 72.1) | 1 (14.3) (0, 37.3) |
| **Male** | 100 (41.2) (35, 47.6) | 53 (36.1) (28.6, 44.2) | 6 (85.7) (71.4, 100) |
| **AGE** |  |  |  |
| **Median Age (Range)** | 45 (9 - 84) | 45 (20 - 84) | 51 (21 - 68) |
| **16 and under** | 1 (0.41) (0, 5.32) | 0 (0) (0, 6.06) | 0 (0) (0, 23.04) |
| **17 to 64** | 202 (83.13) (79.01, 88.04) | 125 (85.03) (80.27, 91.1) | 6 (85.71) (71.43, 100) |
| **65 and over** | 40 (16.46) (12.35, 21.37) | 22 (14.97) (10.2, 21.03) | 1 (14.29) (0, 37.33) |
| **Not reported** | Nil | Nil | Nil |

## Notes

### Competing Interest Statement

The authors have declared no competing interest.

### Funding Statement

No external funding received.

### Author Declarations

Gold Coast Health Human Research Ethics Committee (LNR/2020/QGC/63045)

### Summary of Updates

Minor wording updates and clarifications based on reviewer feedback. - Figure 1: Method for trend line added - Figures 5 & 6: Method for CI and HR added - Max day of repeat test changed from 76 to 72 due to minor error - Tables 1 and supp 1 structure updated for clarity

